# The intersectional role of paternal gender-equitable attitudes and maternal empowerment on child undernutrition in India

**DOI:** 10.1101/2021.02.03.21251084

**Authors:** Anupam Joya Sharma, Malavika Ambale Subramanyam

## Abstract

**Objectives:** Undernutrition is a major global health concern, especially for the low- and middle-income countries. Despite several programmatic efforts by the government, prevalence of undernutrition among Indian children under five years was about 38% in 2015. While studies have extensively investigated the influence of sociodemographic and maternal related factors on child undernutrition, the paternal factors and their interactions with other causal factors are understudied. To fill this gap, we conducted an intersectional analysis to assess the relationship between maternal, paternal, and wealth related factors with stunting and severe stunting among underfives in India.

**Methods:** We used a sample of 22,867 mother-father-child triads from the fourth round of India’s National Family Health Survey (2015-16). We fitted survey adjusted logistic regression models and further adjusted our models for several sociodemographic variables. Results: Our statistical models revealed that even among children from poorer households, those with either an empowered mother or a father with gender-equitable attitudes versus those with none such parents, had a lower odds of stunting (AOR: 0.92, CI: 0.84, 1.02) and severe stunting (AOR: 0.87, CI:0.77, 0.98), independent of all covariates.

**Conclusion:** We argue that while women’s autonomy could reduce the risk of child undernutrition, focusing on men’s attitudes towards gender equity also holds promise for reducing undernutrition. Our findings not only underscore how patriarchy is embodied in undernourished children, but also suggest programmatic interventions to address this deep-rooted scourge in India.

## Introduction

Undernutrition is a major public health problem, causing more than half of deaths of children under five (underfives) globally [1]. However, the burden of undernutrition is higher among the low- and middle-income countries such as India. Despite several programmatic efforts to improve the nutritional status of children in the country, the latest nationally representative survey reported a 38% prevalence of undernutrition among underfives in India [2]. Research from India has highlighted the major role of several factors such as household wealth, maternal education, maternal health in influencing a child’s nutrition status [3]. A growing body of recent studies also suggest a relationship between maternal autonomy and child undernutrition [4]. However, in a patriarchal society such as India, women’s autonomy and access to socioeconomic resources are mediated mostly by their husbands or elderly men in the households [5]. Moreover, such control of women’s movement and behavior by men could lead to low-prioritization of women health in patriarchal households [6]. Since maternal health is strongly associated with child’s health [7], negligence of women’s health could further result in poor child health.

On the other hand, global research has shown that men’s involvement in reproductive and women’s health has a protective effect on women’s and children’s health [8], [9]. However, cultural and social factors in patriarchal societies restrict women’s access to health-promoting resources and inhibit men from participating in decision-making related to women and children [10]. Studies from patriarchal societies such as Bangladesh and Nepal have reported the prevalence of social stigma and embarrassment around men’s involvement in women’s issues [11], [12]. Men’s role in a household is understood as that of a “provider” and “protector” [13], [14]; diverting from these norms by helping one’s wife in cooking and the care of children could result in stigma and sanction by other men [15]. Prevalence of such a socio-cultural stigma suggests a lack of men’s participation in women and childcare in patriarchal societies such as India.

While India-based research on men’s participation in women’s health and childcare is limited, one study from India has shown that men’s knowledge about women’s health and a gender-equitable attitude positively influences maternal health care utilization and also increases women’s autonomy related to their health care [16]. We argue that the husband/partner’s gender-equitable attitudes could not only have a positive effect on the wife’s health, but also result in better nurturing of the child. This suggests that having parents with a woman-friendly outlook—a father with gender-equitable attitudes plus an empowered mother—might lead to better health of the child. However, it is also likely that in a patriarchal society, cultural norms place the wife in a disempowered position despite her husband possessing gender-equitable attitudes. One explanation for this disempowerment in such women could be their internalization of patriarchy [17] despite being married to husbands with gender-equitable attitudes. Similarly, the widespread Indian practice of marital relationships being forged by families and not the two individuals entering matrimony [18] could result in women with a strong sense of independence being married to men who believe in patriarchal traditions. Such marital matches with a mismatch in ideas regarding women’s autonomy might have practical consequences for decision-making related to child rearing, for instance, in matters related to diet or healthcare.

Our study aims to explore the complex relationship of paternal gender-equitable attitudes and maternal empowerment with child nutrition status. We apply the concept of intersectionality [19], [20], a lens which views the social position of an individual within a macro system of intertwining oppressions. This perspective argues that individuals occupy social locations along multiple axes of hierarchy and could simultaneously enjoy a privileged position on one axis and be disadvantaged on another. For instance, a child with a father with gender-inequitable attitudes but with an empowered mother could enjoy the privilege of being nurtured by her mother, but be disadvantaged because of lower levels of paternal nurturing within the house since taking care of the child is conventionally considered as a woman’s responsibility. We hypothesized that paternal gender-equitable attitudes and maternal empowerment are two axes of (in)equality that not only operate together but also interact between themselves in complex ways to affect the nutritional status of their child. Since household wealth has a strong influence on child nutrition status in India [21], we additionally included household wealth as the third axis in our intersectional analysis. The research question addressed in this study was:

How do various intersections of paternal gender-equitable attitudes, maternal empowerment, and household wealth, influence a child’s undernutrition status (stunting) in India?

## Methods

### Data sources and the sample

We draw our data from the fourth round of the Demographic and Health Survey of India (NFHS-4) [2]. The survey was carried out under the supervision of the Ministry of Health and Family Welfare (MoHFW), Government of India with the technical assistance from the United States Inner City Fund (ICF) International, Maryland. Data were collected during 2015 and 2016 from 601,509 households. We merged two files, the children recode (data on children aged 0-5 years) and the couples recode (couple-level data) to generate father, mother, and child triads for our analysis. Anthropometric data were available for 233,695 underfives. However, since information on maternal autonomy and paternal gender-equitable attitudes were not available for a majority of the children, our analytical sample included 22,867 underfives after exclusion of all observations missing data on relevant variables.

### Outcome variables

Stunting or a low height-for-age suggests an inhibition of a child’s linear growth potential and indicates chronic undernutrition. Therefore, we chose stunting and severe stunting as the primary outcomes for our study and assessed them according to the WHO growth standards [22]. A child with a *z* score less than -2 was considered stunted and if below -3, was considered severely stunted.

### Independent variables The intersectional groups

We draw from the concept of women empowerment suggested by Kabeer [23] and operationalized *women’s level of empowerment* using variables indicating women’s participation in decision-making (related to healthcare, household purchases, and visits to friends/relatives and husband’s earnings), perceptions about wife beating (if beating by husband was justified in different situations), ownership of land and house, and current working status (has a job or not). We assigned the value of 1 to each response that indicated the wife’s involvement in decision-making, if the wife disapproved wife-beating, owned any land, owned a house, and if the wife had a job. Upon summing, the values of this measure ranged from 0 to 12. We further dichotomized this measure by splitting at the median (median=8).

*Husband’s gender-equitable attitudes* was operationalized using the variables indicating men’s attitudes towards women’s involvement in decision-making (healthcare and household purchases), whether they thought that it was justified for a wife to refuse sex in different situations and asking him to wear a condom, attitudes towards wife-beating, and attitudes related to contraception. We assigned the value 1 to each response that indicated the husband favored wife’s participation in decision-making, disapproved wife-beating, perceived refusal of sex by wife as justified, perceived the request of condom use by women as justified, and perceived contraception as not just women’s responsibility. Upon summing, the measure had values ranging from 0 to 16. We then dichotomized the measure by splitting at the median (median=14).

A household wealth index was created using information on the ownership of several consumer goods and household characteristics, such as availability of clean drinking water, ownership of television and so on. [2]. The dataset included information on household wealth quintiles (richest, richer, rich, poor, and poorest) based on these scores. However, we merged the richer and richest quintiles to represent the affluent; and the rich, poorer, poorest to represent the non-affluent categories.

Six intersectional groups were created representing the intersection of the three axes:

1. Concordant-high and affluent group: Children having a father with high gender-equitable attitudes, a mother with high levels of women empowerment (labeled “high” because we assumed that this pairing represents greater woman-friendly environment) and belonging to an affluent household.
2. Concordant-high and non-affluent group: Children having a father with high gender-equitable attitudes, a mother with high levels of women empowerment, and belonging to a non-affluent household.
3. Discordant affluent group: Children having a father with high gender-equitable attitudes but a mother with lower levels of women empowerment *or* having a father with low gender-equitable attitudes but an empowered mother *and* belonging to an affluent household.
4. Discordant non-affluent group: —Children having a father with high gender-equitable attitudes but a mother with lower levels of women empowerment or having a father with low gender-equitable attitudes but an empowered mother and belonging to a non-affluent household.
5. Concordant-low and affluent group: —Children having a father with lower gender-equitable attitudes, a mother with low levels of women empowerment (labeled “low” because we assumed that this pairing represents low levels of woman-friendly environment) and belonging to an affluent household.
6. Concordant-low and non-affluent group—Children having a father with lower gender-equitable attitudes, a mother with low levels of women empowerment, and belonging to a non-affluent household.

### Sociodemographic indicators

The covariates included in our analysis were father’s total years of education, mother’s total years of education, religion and caste of the parents, age of the father (in years), age of the mother (in years), place of residence (rural/urban), gender of the child (male/female), birth order of the child, and mother’s body mass index.

### Statistical analyses

We first calculated the prevalence of stunting and severe stunting among the underfives across all intersectional groups. Later, we fitted survey-design adjusted logistic regression models to estimate the association of the six intersectional groups with the outcomes--stunting and severe stunting. We added the following covariates to our logistic models (separate for each outcome): father’s education, mother’s education, religion, caste, age of the father, age of the mother, place of residence, gender of the child, birth order of the child, and maternal body mass index. Our models also accounted for the clustering due to the sampling design and the women’s sample weight. *Alpha* was set at 0.05.

All survey-adjusted logistic regression models were run in Stata version 12 [24].

### Ethical considerations

We used de□identified Demographic and Health Survey data for India from 2015-16 [2]. The survey had ethical approval from the institutional review board of International Institute of Population Sciences, Mumbai, India. Written informed consent was taken from all adult participants. In the case of minors, consent was taken from their parents.

### Patient and Public Involvement

No patients or public were involved in this research. All analyses was based on publicly available secondary data.

## Results

### Descriptive results

The mean maternal age in our sample was 27.42 years (SD=5.12), mean paternal age was 31.86 years (SD=6), and mean child age was 2.04 years (SD=1.40). About 52% of the children in our sample were male and a majority (∼60%) were first or second in birth order. About 74% of the mother-father-child triads were urban dwellers and about 68% belonged to poorer households. The prevalence of stunting and severe stunting was considerably low across the affluent intersectional groups than among groups that were non-affluent (Table 1). Within a given affluence category, concordant-high and discordant groups had a lower prevalence of stunting (and severe stunting) versus concordant-low groups.

**Table 1:** The distribution of stunting and severe stunting (%) among Indian underfives across the six intersectional groups (N=22,867).

| Intersectional groups | Stunting |  | Severe stunting |  |
| --- | --- | --- | --- | --- |
|  | N | Prevalence (%) | N | Prevalence (%) |
| Concordant-high and non-affluent group | 1,118 | 41.65 | 457 | 17.03 |
| Concordant-high and affluent group | 452 | 24.24 | 140 | 7.51 |
| Discordant non-affluent group | 3,182 | 43.09 | 1,331 | 18.03 |
| Discordant affluent group | 984 | 26.23 | 359 | 9.57 |
| Concordant-low and non-affluent group | 2,774 | 46.17 | 1,249 | 20.79 |
| Concordant-low and affluent group | 661 | 27.18 | 268 | 11.02 |

### Results from intersectional analysis

Results from survey adjusted logistic regression models showed that children belonging to the concordant-high and affluent group had the lowest odds (AOR:0.56, CI:0.46, 0.67) of stunting compared to those belonging to concordant-low and non-affluent group (Table 2), independent of their father’s education, mother’s education, religion, caste, age of the father, age of the mother, place of residence, gender of the child, birth order of the child, and maternal body mass index. We also found that independent of all covariates, children from the discordant non-affluent group, compared to the concordant-low and non-affluent group, had lower odds of stunting (AOR: 0.92, CI: 0.84, 1.02) and severe stunting (AOR: 0.87, CI:0.77, 0.98). Findings for severe stunting showed one notable pattern: Substantially lower odds of severe stunting in both the groups with parental concordance in woman-friendly outlook, whether affluent or non-affluent.

**Table 2:**
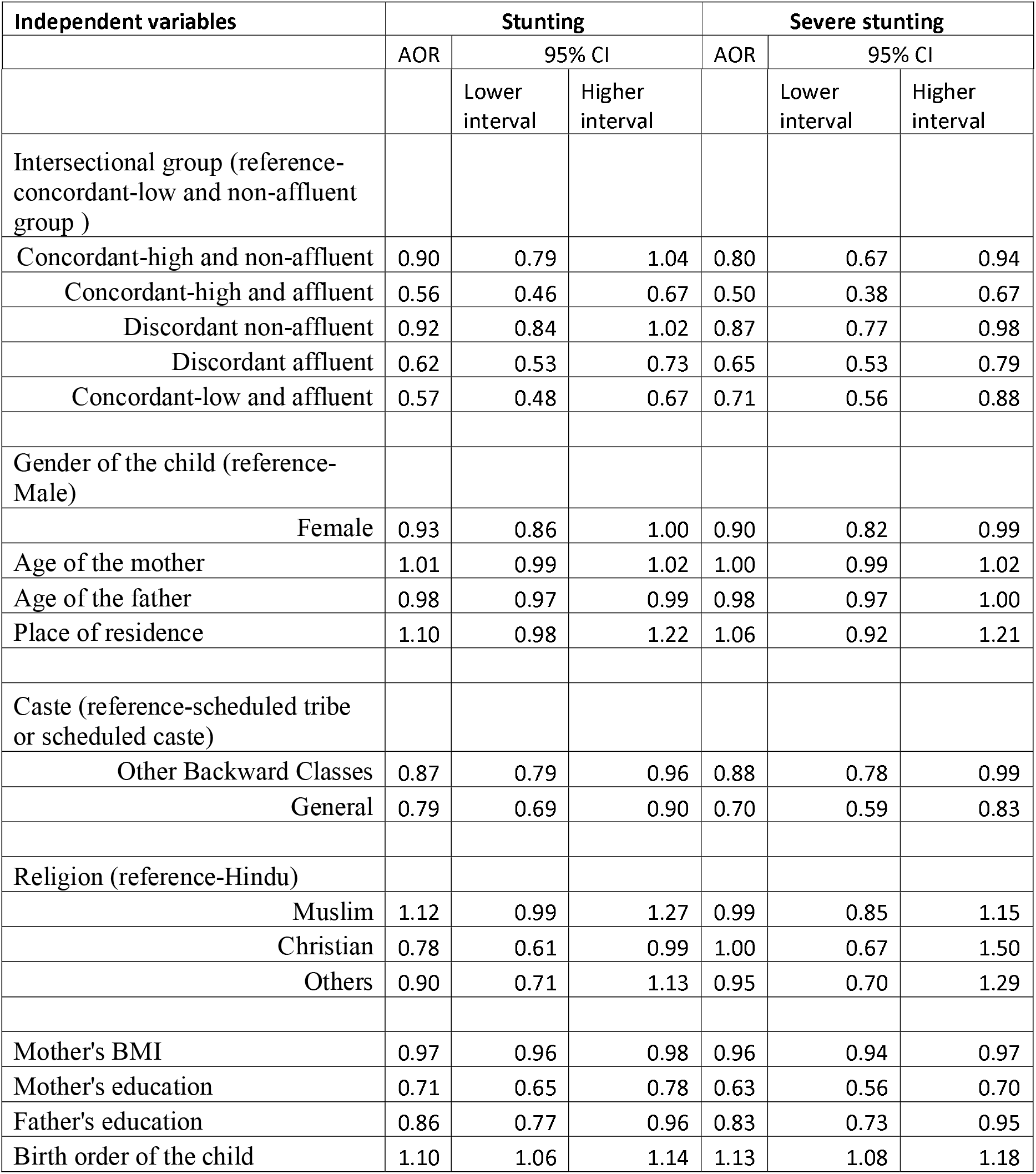
Association (adjusted odds ratios, 95% confidence interval (CI)) of the groups representing the intersection of maternal autonomy, paternal gender-equitable attitudes, and household wealth with stunting and severe stunting among underfives in India (N=22,867).

A greater maternal body mass index (BMI), more years of mother’s and father’s education, and a lower birth order of the child were associated with lower odds of stunting and severe stunting in our adjusted logistic regress models.

## Discussion

Our intersectional analysis using a large community-based sample of parents-child triads suggests that being raised by parents who are more woman-friendly could lower the risk of their child being undernourished compared to being raised by less woman-friendly parents. Previous research from India has primarily focused on the influence of maternal autonomy on child nutrition [4]. While a husband’s gender-equitable attitudes may have been shown to influence his wife’s empowerment [16], we argue that it could also be directly and indirectly related to their child’s wellbeing. However, we could not locate research from India that investigated the relationship of paternal gender related attitudes with child undernutrition.

We found that irrespective of the wealth of the household, children having either an empowered mother or a father with higher gender-equitable attitudes had lower odds of stunting and severe stunting compared to children of poorer households with a disempowered mother and a father with lower gender-inequitable attitudes. This suggests that having at least one parent who could create a woman-friendly environment for the child in the household might improve the nutrition status of the child. The importance of having a parent, any parent, with a woman-friendly outlook is thus underscored and the supremacy of the influence of maternal, compared to paternal, characteristics on child health is questioned.

Arguably, an empowered mother could have a greater say in household decision-making, thus accessing the resources to take better care of her child. For instance, she could take decisions related to her own health (which could affect the health of the child), what should be cooked in the house (which could affect the child’s nutrition status), and decisions directly related to the child’s healthcare. This is corroborated by previous research from South Asia highlighting the relationship between women’s autonomy and child undernutrition [25]. On the other hand, a father with greater gender-equitable attitudes could be more flexible with gender-roles within the household, thereby being more active in childcare or helping the mother with household chores. The nurture and care from the father may reduce the risk of child undernutrition. Previous studies from patriarchal societies such as Malawi have highlighted the benefits of men’s involvement in maternal and child health for improved nutritional status [26]. While such paternal involvement in childcare is likely uncommonly observed in patriarchal societies such as India, a previous study from Pakistan suggested that paternal involvement in childcare is crucial for child wellbeing [27]. The implication then is that among other benefits, children’s health will likely improve through efforts to instil more gender equitable attitudes among men. Indian men’s gender-equitable attitudes were found to improve their involvement in women’s healthcare [16], and also improve women’s autonomy related to household decision making in India [16], which in turn might lead to better child health [4]. While these explanations and learnings from previous studies discuss how maternal and paternal influences, independently, may impact a child’s nutritional status, our study was able to highlight that when examined from an intersectional perspective, the child appears to benefit equally from having *a* parent with a woman-friendly perspective, regardless of the parent’s gender. This highlights the contribution of the father’s attitude in a realm which is traditionally considered to be influenced heavily by maternal factors.

Moreover, our intersectional analyses allowed us to show that simultaneously occupying a privileged location on two axes (having an empowered mother and a father with a greater gender-equitable attitudes) substantially reduced the odds of child undernutrition, compared to having only either/none such parents. This would be missed in most analyses that estimate the role of maternal and paternal factors *independent* of each other. A high woman-friendly parental unit likely offers the child greater opportunities to receive nurturing from both the parents and benefit from both maternal and paternal resources for overall wellbeing, thereby reducing the risk of undernutrition. It is therefore not surprising that children of mothers who did not report greater empowerment and fathers with lower gender-equitable attitudes had the greatest odds of undernutrition.

The strong role played by household wealth was highlighted in our findings. Children belonging to richer households had lower odds of undernutrition compared to those from poorer households, for every combination of maternal empowerment and paternal gender-equitable norms. Greater economic resources could create a healthier environment for the child through several pathways, including via increased access to a healthier diet and ability of parents meet the demands of healthcare expenditure. While previous studies from India have shown that children belonging to poorer versus richer households were at greater risk of undernutrition [21], [28], our intersectional analysis revealed interesting results. We found that children belonging to poorer households but having either an empowered mother or a father with a greater gender-equitable attitudes had lower odds of undernutrition compared to children of poorer households and having none such parents. This suggests that despite a lack of economic resources in the household, having an empowered mother or a father with gender-equitable attitudes could act as an unmaterialistic yet invaluable resource promoting the child’s health. Previous studies have shown that the need and demand for childcare in low-income families are not different from families with economic privileges, however, they may lack choices due to their pressing need to work more to sustain their family [29].

However, having an empowered mother or a father having greater gender-equitable attitudes in such households could result in a flexible division of household labour thus creating more ways to negotiate their daily work schedules and devote more time to childcare. Moreover, programs and policies focused on reducing undernutrition would likely target the poor due to the persistent wealth inequalities in undernutrition in India [30]. An empowered mother or a father with greater gender-equitable attitudes could be more receptive to the messages from, and more conscientious about accessing the benefits of, these programs due to their open outlook, thereby reducing the risk of their child’s undernutrition. For instance, a father with greater gender-equitable attitudes could be more attentive of maternal healthcare related information and actively participate in community events organized locally. Similarly, an empowered mother could have more frequent interaction with the community health workers such as the Accredited Social Health Activists and Anganwadi workers about her own and her child’s health. Such empowered mothers might also be more comfortable participating in community events due to support from husbands with greater gender-equitable attitudes.

Overall, our findings suggest that children with a privileged position in any one of the three intersectional axes (maternal autonomy, paternal gender-equitable attitudes, and household wealth) were better off compared to the most disadvantaged children. These children could offset the effects of their disadvantaged position through the privileges gained from the other axes.

## Limitations and strengths

The study has several limitations that need to be considered. First, the cross-sectional nature of the survey limits our ability to draw causal conclusions. However, the statistically significant associations provide evidence to support the interesting relationship between the intersectional groups and undernutrition among Indian underfives. Second, despite the larger sample of children with anthropometric data we could use only a subsample which had data on gender-related attitudes of the parents, thereby limiting our ability to generalize the findings. Our sample also comprised respondents majorly from urban areas. However, our sample had a good diversity of social groups such as caste, religion, gender, and educational attainment that allowed us to make several interesting conclusions. Lastly, due to the smaller sample size we could not explore the intersection of other social axes such as caste, place of residence, and so on, which could have yielded a more comprehensive picture. Additionally, due to low statistical power we could not assess the relationship of discordant-paternal high and discordant-maternal high groups with the outcomes, separately. Nevertheless, we argue that despite the modest sample size we found statistically significant results in our analysis that contributes to an understudied topic. Furthermore, the findings from our analytical models that accounted for several important social factors shed light on the importance of women-friendly parental characteristics in improving child health.

Despite these limitations, to our knowledge, this is the first study to apply the lens of intersectionality to explore the relationship of parental characteristics regarding gender-roles and household wealth with child nutritional status thereby creating new avenues in understanding child undernutrition in India.

## Implications and conclusion

In this paper, we argue that while women’s autonomy potentially reduces the risk of child undernutrition, focusing on men’s attitudes towards gender equity also holds promise for improvement in child health, especially undernutrition. Therefore, these findings are relevant to the design and reform of a variety of policies such as those that focus on improving women’s social status in society, changing men’s views about gender roles to be more egalitarian, and reducing wealth-inequity among Indian households. More specifically, our results emphasize that men’s involvement in child health could be integrated into existing programs targeting undernutrition, such as the Integrated Child Development Services.

Interventions related to paternal involvement in child-care could also be introduced. Local non-governmental organizations could play an active role in the design and demonstration of community-based programs that simultaneously target women empowerment and men’s gender-related attitudes. For instance, previous research highlighted that men’s involvement in maternal and childcare through policies and interventions changed the perception of men about gender-roles and dynamics at the community level [26]. Finally, our findings open avenues for researchers to conduct further research, especially longitudinal investigation, focused on unpacking the complex process linking parental gender-related perspectives and child undernutrition.

The intersectional lens has allowed us to highlight the impact on child health of a father’s gender-equitable attitude despite his wife not being highly empowered. The widespread practice of focusing on maternal characteristics, while occasionally accounting for paternal factors, would miss such insights. Our findings not only underscore how patriarchy is embodied in undernourished children, but also suggest programmatic interventions to address this deep-rooted scourge of India.

## Data Availability

All relevant data are publicly available in the DHS website.

## Contributorship statement

Anupam Joya Sharma: Conceptualization, Formal analysis, Investigation, Methodology, Project administration, Writing – original draft

Malavika Ambale Subramanyam: Conceptualization, Project administration, Supervision, Writing – review & editing

## Competing interest

There are not competing interests to state.

## Funding

The study has received no funding.

## Data sharing statement

The data used in this study is publicly available at the DHS website (https://dhsprogram.com/methodology/survey-Types/dHs.cfm).

## Data availability

The datasets generated during and/or analysed during the current study are available in the DHS repository: https://dhsprogram.com/what-we-do/survey/survey-display-541.cfm

## Notes

### Competing Interest Statement

The authors have declared no competing interest.

### Funding Statement

No external funding was received for this project.

### Author Declarations

Institutional Ethics Committee, Indian Institute of Technology Gandhinagar, India

